# ViViEchoformer: Deep Video Regressor Predicting Ejection Fraction

**DOI:** 10.1101/2024.06.21.24309327

**Authors:** Taymaz Akan, Sait Alp, Md. Shenuarin Bhuiyan, Tarek Helmy, A. Wayne Orr, Md. Mostafizur Rahman Bhuiyan, Steven A. Conrad, John A. Vanchiere, Christopher G. Kevil, Mohammad A. N Bhuiyan

**Affiliations:** Department of Medicine, Louisiana State University Health Sciences Center at Shreveport, Shreveport, LA 71103, USA; Department of Computer Engineering, Erzurum Technical University, Erzurum, Turkey; Department of Pathology and Translational Pathobiology, Louisiana State University Health Sciences Center at Shreveport, Shreveport, LA 71103, USA; Department of Molecular and Cellular Physiology, Louisiana State University Health Sciences Center at Shreveport, Shreveport, LA 71103, USA; Department of Pediatric Cardiology, Bangabandhu Sheikh Mujib Medical University, Bangladesh; Department of Pediatrics, Louisiana State University Health Sciences Center at Shreveport, Shreveport, LA 71103, USA

## Abstract

Heart disease is the leading cause of death worldwide, and cardiac function as measured by ejection fraction (EF) is an important determinant of outcomes, making accurate measurement a critical parameter in PT evaluation. Echocardiograms are commonly used for measuring EF, but human interpretation has limitations in terms of intra- and inter-observer (or reader) variance. Deep learning (DL) has driven a resurgence in machine learning, leading to advancements in medical applications. We introduce the ViViEchoformer DL approach, which uses a video vision transformer to directly regress the left ventricular function (LVEF) from echocardiogram videos. The study used a dataset of 10,030 apical-4-chamber echocardiography videos from patients at Stanford University Hospital. The model accurately captures spatial information and preserves inter-frame relationships by extracting spatiotemporal tokens from video input, allowing for accurate, fully automatic EF predictions that aid human assessment and analysis. The ViViEchoformer’s prediction of ejection fraction has a mean absolute error of 6.14%, a root mean squared error of 8.4%, a mean squared log error of 0.04, and an *R*^2^ of 0.55. ViViEchoformer predicted heart failure with reduced ejection fraction (HFrEF) with an area under the curve of 0.83 and a classification accuracy of 87 using a standard threshold of less than 50% ejection fraction. Our video-based method provides precise left ventricular function quantification, offering a reliable alternative to human evaluation and establishing a fundamental basis for echocardiogram interpretation.

## INTRODUCTION

Cardiovascular diseases (CVDs) encompass a range of conditions that can negatively impact the health of the cardiovascular system, which consists of the heart and blood vessels. CVDs are consistently ranked as one of the top causes of death worldwide ^1^. Heart failure (HF) is a rapidly growing cardiovascular condition, with an estimated prevalence of 37.7 million individuals worldwide. HF is a chronic phase of cardiac functional impairment, causing symptoms such as dyspnea, fatigue, poor exercise tolerance, and fluid retention, which impact patients’ quality of life and contribute to the global health crisis ^2^. It also carries a high mortality rate. Diagnosing heart failure requires an accurate assessment of cardiac function, which can be done using various methodologies to quantify and characterize. Left ventricular EF is one of the most important metrics for assessing cardiac function, which measures how well the left ventricle can eject blood ^3,4^.

Standard methods for estimating left ventricular ejection fraction include echocardiograms, cardiac MRI, cardiac computed tomography (CT), and Equilibrium Radionuclide Angiocardiography (ERNA). Echocardiography uses ultrasound to create real-time images of the heart’s chambers, valves, and blood flow, assessing the volume of blood pumped out of the left ventricle with each contraction. MRI provides detailed images of the heart’s structure and function but has limitations such as cost, availability, and potential contraindications. CT uses X-rays to produce detailed heart images but has limitations such as radiation exposure, allergic reactions to contrast media, and limited dynamic heart function assessment.

Equilibrium radionuclide angiography is a method used in nuclear medicine studies. Still, it has some drawbacks, like taking a long time to process, injecting radiopharmaceutical agents, and yielding low resolution for regional ventricular function in heart disease patients ^3,5^. Clinically echocardiography is the preferred most common method for estimating LVEF because it is widely available, provides real-time imaging, is non-invasive, and is more cost-effective than other options. This makes it particularly useful for quick and detailed assessments in various clinical situations.

Traditional echocardiography typically includes a visual interpretation to estimate LVEF, providing a qualitative assessment without precise numerical values. This approach is well-suited for managing acute patients but falls short when it comes to serial evaluations, particularly in patients with valvular lesions causing regurgitation. There are also quantitative capabilities for echocardiography using the Simpson”s method and fractional shortening to calculate EF. The human calculation of ejection fraction is subject to variability due to irregular heart rate and the nature of the calculation, which necessitates manual ventricle size tracing for every beat ^4^. The variability in estimating LVEF among different observers can often result in requests for additional testing, review of the study, and reinterpretation, which can impact the timing of therapeutic interventions ^5–7^.

Conventional Machine learning (ML) has recently led to substantial advancements in diverse fields, including medical applications. Conventional ML has been utilized in echocardiography to determine the ejection fraction, with significant interest in their potential to provide improvement in disease diagnoses, aid decision-making, and serve as a confirmatory assessment ^8,9^. However, conventional ML has a potential disadvantage in its reliance on feature engineering, which is a manual and time-consuming process. Moreover, despite obtaining images in various positions and orientations, these conventional echocardiographic systems lack 3D localization and spatial relation measurements for volume computation.

Deep learning has driven a significant resurgence in machine learning due to availability of large data sets, and advances in computing power ^10–15^. This field has revolutionized machine learning by understanding and manipulating data, including images ^16,17^, and incorporation of natural language processing (NLP) ^18^. Moreover, deep learning differs from conventional methods as it avoids manual feature engineering. Also, using deep learning techniques in medical diagnostics improves the accuracy of diagnoses ^19,20^. It plays a crucial role in predictive analytics, allowing for detecting possible health risks or outcomes ^21^. These techniques provide healthcare professionals with valuable predictive insights through the assimilation and analysis of various datasets, including patient information, genetic profiles, imaging studies, and clinical records, and enables early detection of diseases or health deterioration ^22^.

Deep learning techniques can be used to determine ejection fraction, estimate end-diastole and end-systole volumes, and calculate the percentage difference between them, rather than relying on actual echocardiogram videos ^23–27^. A recently proposed method, EchoNet-Dynamic ^4^, directly regresses LVEF from video inputs using spatiotemporal models, which avoids the need to estimate EDV and ESV separately. EchoNet-Dynamic, a video-based deep learning model, has been proposed for echocardiograms, demonstrating its ability to assess ejection fraction accurately across the entire video. It is a CNN model that uses atrous convolution ^28^ for semantic segmentation of the left ventricle, a CNN model ^29^ with residual connections and spatiotemporal convolutions for predicting the ejection fraction, and video-level predictions for beat-to-beat estimations of cardiac function. Moreover, another video-based method performs LV segmentation using echocardiogram sequences and then converts the predicted context into an end-to-end video regression model ^30^. However, segmentation, a sensitive process involving categorizing entire regions, may increase computational requirements and processing times. Inaccuracies in segmentation can impact subsequent classification or regression tasks, making the overall process more sensitive to segmentation quality. Recent advances in deep learning have shown that it can accurately and reproducibly identify human-identifiable phenotypes and characteristics not recognized by human experts, overcoming limitations in human interpretation ^31–33^.

Herein, we propose an end-to-end deep learning approach, ViViEchoformer, which leverages a video vision transformer (ViViT) ^34^ to regress LVEF from echocardiogram videos directly. We converted ViViT from classification to regression to predict LVEF. The model captures spatial information and preserves inter-frame relationships by extracting spatiotemporal tokens from the input video. While utilizing the video vision transformer to capture spatiotemporal patterns in the video accurately, this method performs precise, fully automatic EF predictions that facilitate human assessment and subsequent analysis.

## RESULTS

Our neural network architecture was implemented in Python using the TensorFlow and Keras libraries. A workstation equipped with 62 GB of RAM and an NVIDIA GeForce GTX 4080 GPU was used for all experiments. We trained our transformer model (**Fig 1**) on a data set with 10,030 echocardiogram videos provided by Stanford University Hospital ^35^. We converted the classification model ViViT, into a regression model and trained it to estimate the left ventricular ejection fraction from echocardiogram videos using a training and validation set of over 30700 and 1200 videos, respectively, and a test set of over 1200 videos. The analysis focused on the 32 frames of videos that were resized to 52×52 dimensions.

**Fig 1.**
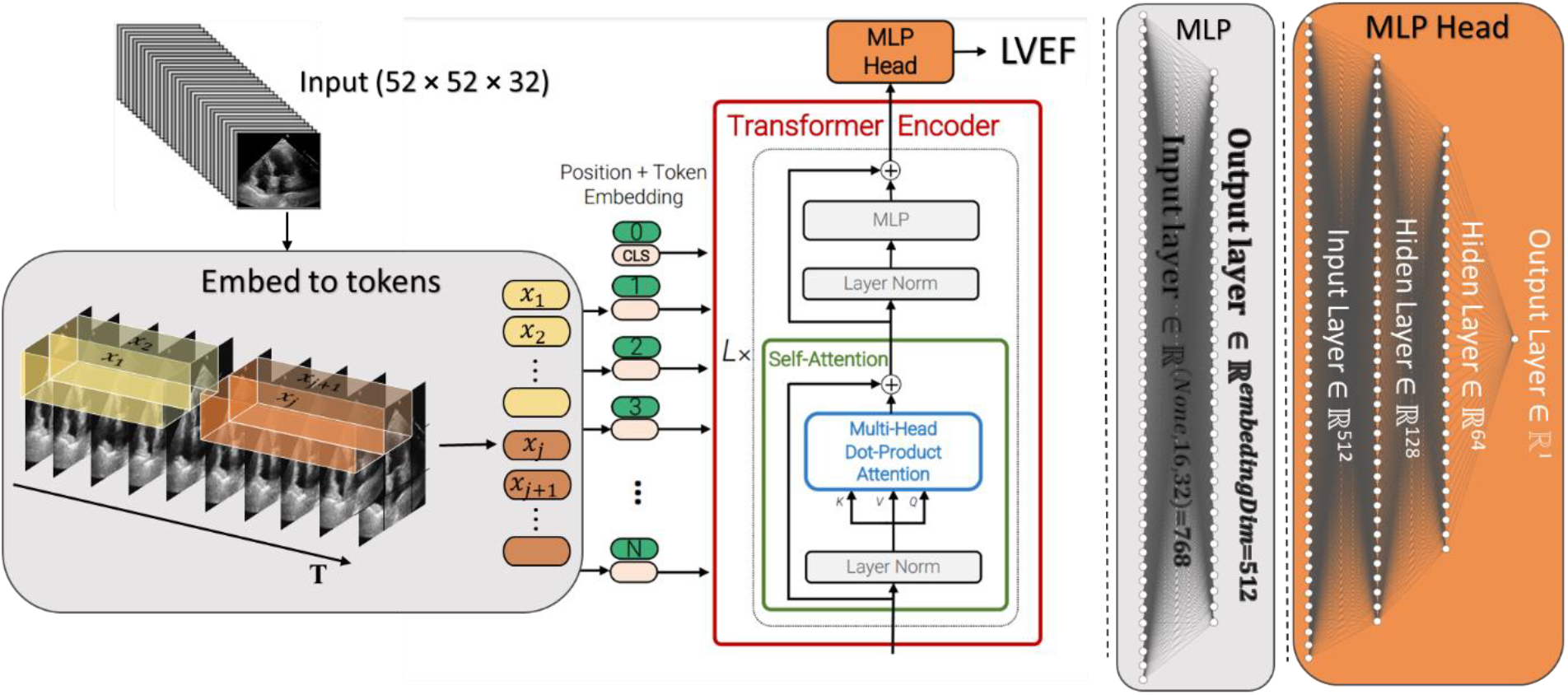
The model pipeline for video regression. The Tubelet embedding technique extracts and linearly embeds nonoverlapping tubelets across the spatio-temporal input volume. Using spatial-temporal attention, the transformer encoder forwards all spatio-temporal tokens extracted from the video.

We employed the SGD optimizer for training, and the training process was conducted over 100 epochs with a batch size of 128 and a learning rate of 1e-4. **Table 1** provides a summary of the configuration of the training parameters. The model checkpoint is configured to save only the optimal solution discovered during training based on the loss function evaluation during validation. The model checkpoint is saved when a metric improves on a validation set during training. As depicted in **Fig 2**, the model demonstrates a significant reduction in loss in the initial epochs, which indicates the model’s capacity to learn quickly from the training data.

**Table 1.** Details of model variants.

| Parameter name | Values |
| --- | --- |
| **Hyperparameters of ViViT** |  |
| Optimizer | SGD |
| Batch size | 128 |
| Epoch | 100 |
| Input Shape | (52, 52, 32, 1) |
| Layer norm | 1e-6 |
| Learning rate | 1e-4 |
| Number of heads | 12 |
| Number of Layers | 10 |
| Patch size | (32, 8, 8) |
| Projection dim | 512 |

**Fig 2.**
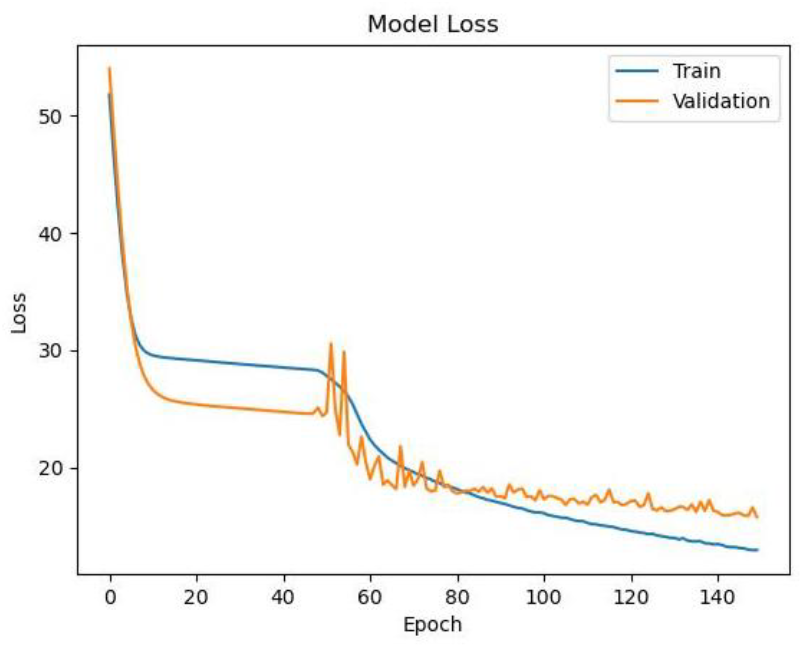
The graph illustrates the model’s loss over epochs for training (blue) and validation (orange) datasets.

We have employed the evaluation metrics for evaluating the performance of ViViEchoformer on the EchoNet test dataset, which were not previously used during model training. The estimation of EF has been associated with interobserver variability of up to 14% ^36^. The ViViEchoformer’s prediction of ejection fraction had a mean absolute error of 6.14%, root mean squared error of 8.4%, mean squared log error of 0.04, and an *R*^2^ of 0.55.

The visual assessment has been carried out using six plots (**Fig 3**). These plots are used to evaluate the performance of a predictive model, providing information about the accuracy, distribution of errors, and independence of errors, which are crucial for validating the robustness of the model. The scatter plot **Fig 3**a shows the model’s predictions against the actual values, with points scattered around the line of perfect agreement. This indicates that the model captures the trend in the data, but the spread of points away from the line indicates variances in prediction accuracy. The violin plot and histogram of error distribution **Fig 3**b, c provide insight into the distribution of prediction errors, with a long tail of errors indicating a right-skewed distribution. The line plot of errors in **Fig 3**d shows variability, with most falling within two standard deviations of the mean. However, occasional spikes beyond this range suggest more significant errors, possibly due to outliers or less valid assumptions. The autocorrelation and partial autocorrelation plots in **Fig 3**e, f show that the errors are mostly independent, indicating a positive predictive model performance.

**Fig 3.**
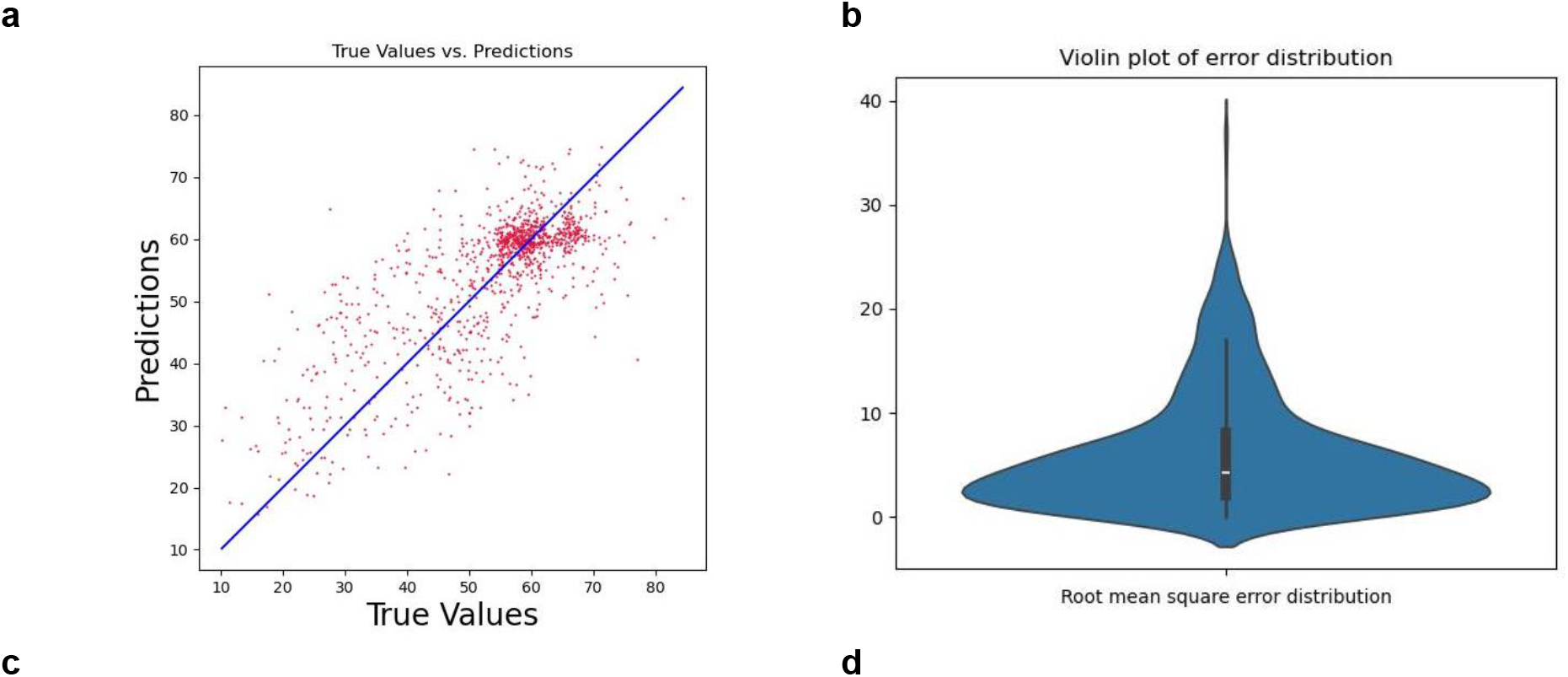
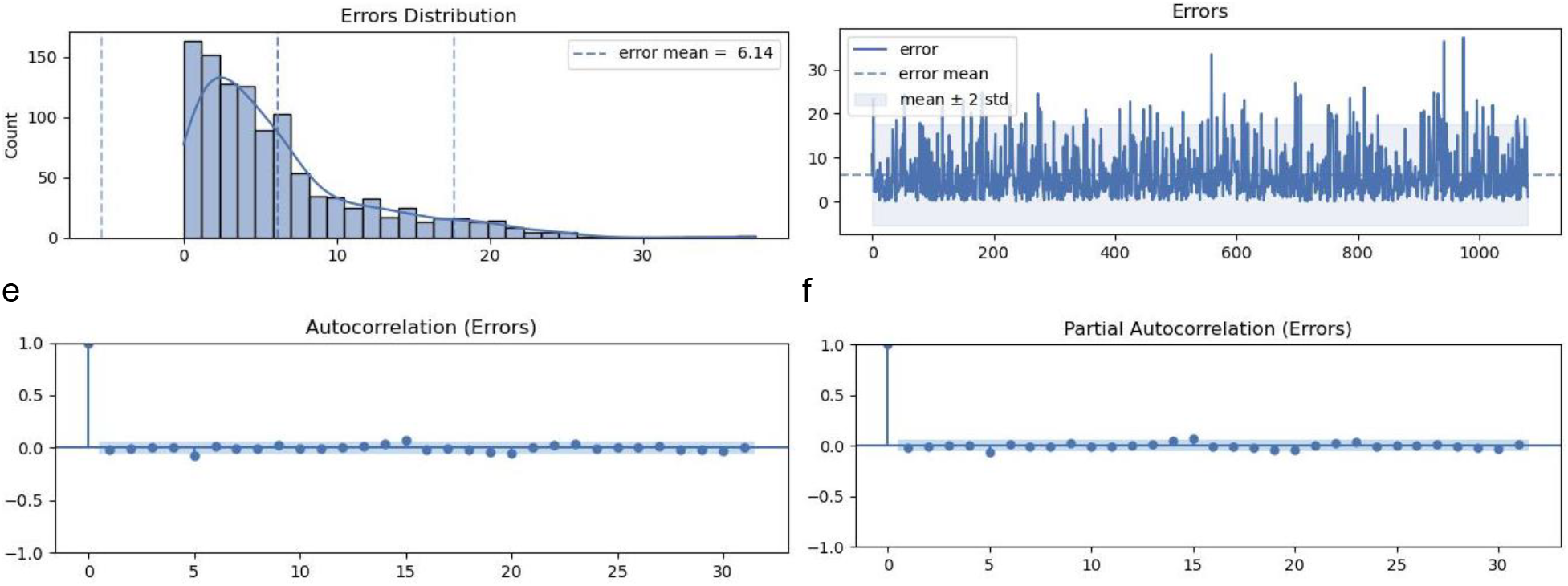
Model Evaluation. **a** Comparison of ViViEchoformer predicted, and EchoNet dataset reported ejection fractions (*n* = 1288). **b** the violin plot showcasing the model error distribution. **c** errors distribution histogram. **d** error values across samples. **e** Autocorrelation plot of residuals. **f** partial autocorrelation of the residuals.

**Table 2** reports the model’s classification performance distinguishing between Heart Failure with Reduced Ejection Fraction (HFrEF) and Non-HFrEF cases. Precision, recall, f1-score, and support numbers are reported for both categories. The classification report shows ViViEchoformer’s prediction of HFrEF with an area under the curve of 0.83 (**Fig 4**a), using a common threshold of an EF of less than 50%. The model achieves a precision of 0.77 for HFrEF cases, indicating 77% correctness, and a recall of 0.83, indicating 83% correct identification. The f1-score balances these metrics, indicating the model’s effectiveness in HFrEF cases. However, the model performs better for non-HFrEF cases, with a precision of 0.91 and recall of 0.92, resulting in a higher f1-score of 0.89. The overall accuracy across both classes is 0.87, indicating 87% correct classifications. The macro average f1-score is 0.83, considering the balance between classes without weighting for their representation in the dataset. The weighted average f1-score is also 0.87, indicating consistent high performance across classes when accounting for the number of samples in each. **Fig 4**b illustrates the confusion matrix for our model’s classification performance where HFrEF is labeled 0, and Non-HFrEF is labeled 1. The matrix visually represents the model’s predictions compared to the actual labels.

**Table 2.**
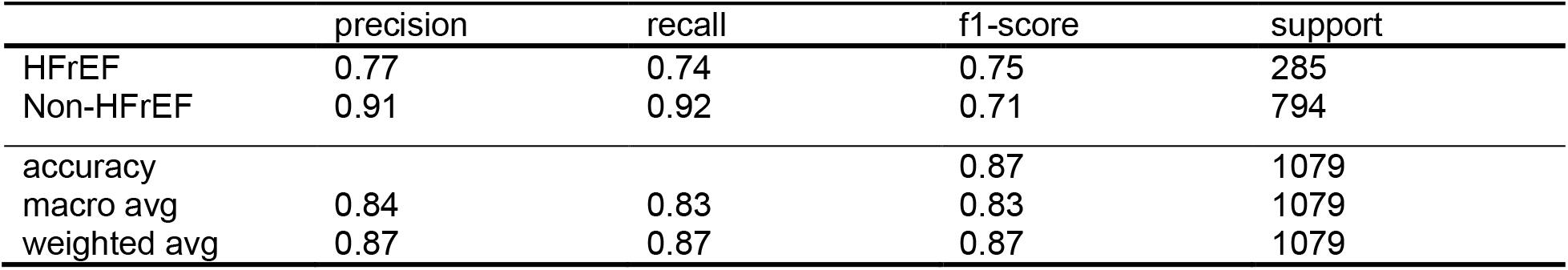
Classification performance for HFrEF and Non-HFrEF cases.

**Fig 4.**
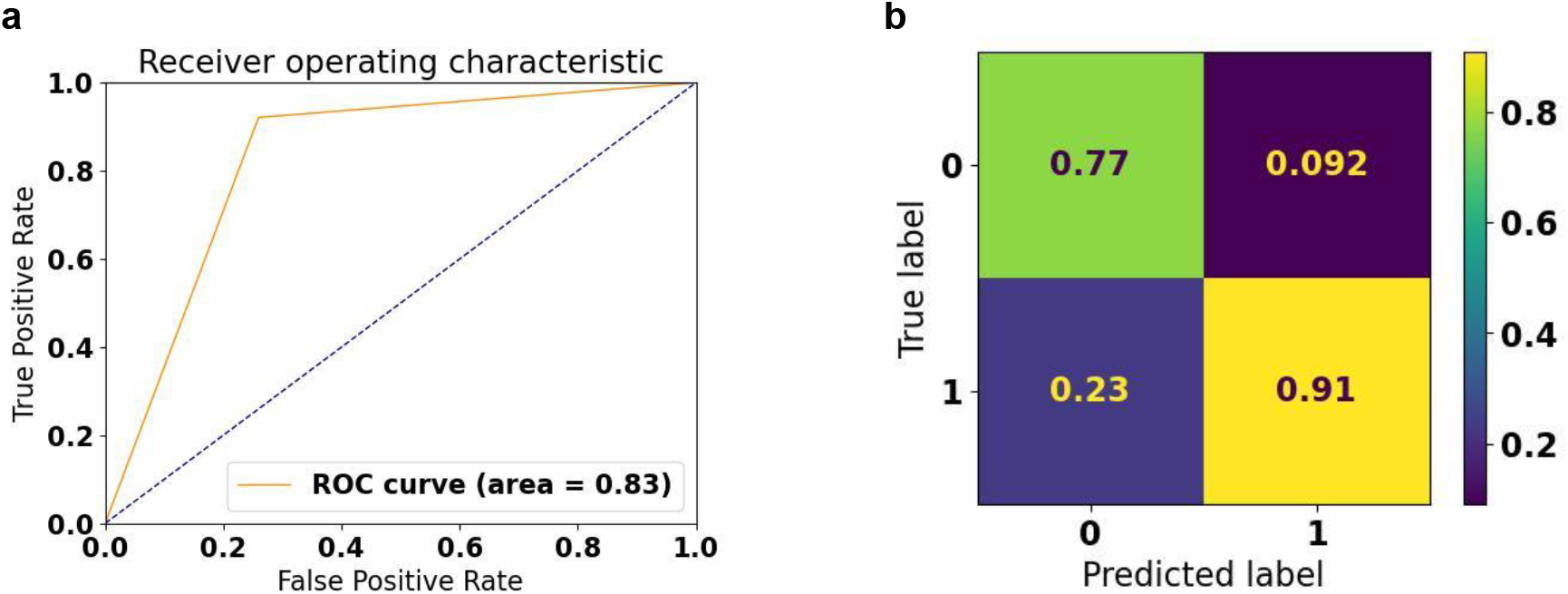
Classification accuracy of 85 and AUC of 0.83 using a standard threshold of less than 50% EF. a ROC curve, b Confusion matrix where HFrEF, labeled as 0 and non-HFrEF labeled 1.

## DISCUSSION

The most prominent architecture of choice in sequence modeling is the transformer, which uses a multi-headed self-attention mechanism instead of convolution. ViViEchoformer is a video transformer-based deep learning model for echocardiogram video understanding tasks, allowing for accurate, fully automatic EF predictions that aid human assessment and analysis. To our knowledge, ViViEchoformer is the first deep-learning model that uses transformers to estimate the ejection fraction from echocardiogram videos. Previous attempts to use deep learning techniques are primarily used to determine EFs, end-diastole and end-systole volumes, and percentage differences in echocardiogram videos rather than actual data. These methods typically do not account for inter-frame relationships or temporal dependencies within the video sequences during their analysis. To process video sequences, ViViEchoformer splits them up into smaller temporal and spatial units known as tokens. The model can then comprehend temporal dependencies throughout the sequence and spatial relationships within individual frames thanks to extracting and processing information from these tokens across frames.

Some video-based methods perform LV segmentation using echocardiogram sequences, but segmentation may increase computational requirements and processing times due to its sensitive nature. However, when analyzing massive datasets, DL techniques can reveal hidden patterns that were previously not apparent. Recent advancements in DL techniques have demonstrated their ability to “see the unseen” in images and videos. Consequentially, determining EFs without end-diastole and end-systole volumes could be possible for DL techniques. Without infusing knowledge awareness and using any pre-processing, such as segmentation, our method directly regresses EF among the video frames. ViViEchoformer’s predictions have a variance comparable to or less than human experts’ measurements of cardiac function ^37^. ViViEchoformer achieved high prediction accuracy for estimating ejection fraction performed by human interpreters. Its prediction of ejection fraction had a mean absolute error of 6.14%, which is within the typical inter-observer variation of 14%.

In the study by Ouyang et al. ^4^, five expert sonographers and cardiologists conducted a blinded review of echocardiogram videos that exhibited the largest absolute differences between the initial human labels and the predictions made by EchoNet-Dynamic. These experts independently assessed the relevant videos and two blinded measurements of ejection fraction. The findings revealed that 38% (15 out of 40) of the videos had significant issues related to video quality or the acquisition process. In comparison, 13% (5 out of 40) were characterized by marked arrhythmias, which constrained the experts’ capacity to assess ejection fraction accurately. A critical limitation of the EchoNet-Dynamic dataset stems from the inaccuracy in the initial human labeling of echocardiogram videos, compounded by issues related to poor image quality, arrhythmias, and variations in heart rate. These factors significantly impact on the training and evaluation performance of our model.

In developing a model to regress the left ventricular ejection fraction (LVEF) from echocardiogram videos, we encountered a nuanced issue at the intersection of statistical significance and clinical utility, particularly when classifying LVEF based on the 50% cutoff. Our model is capable of closely approximating actual LVEF values. Yet, we observe instances where minor discrepancies—such as a predicted LVEF of 49.9% versus an actual measurement of 50.01%—raise important considerations. While these small differences may be statistically significant, they highlight the clinical uncertainty of near-threshold predictions in model evaluation. This distinction is important because, in clinical practice, the marginal difference may not change treatment or patient outcome, calling statistically significant but clinically marginal model predictions into question. This is a limitation for most methodologies and should be acknowledged.

**Fig 5** presents a scatter plot evaluating the performance of a regression model that predicts left ventricular ejection fraction (LVEF). The true LVEF values are on the X-axis, while the Y-axis displays the model’s predicted LVEF values. The overlay of a green zone and an orange area indicates the boundary of correct and incorrect classifications by the model relative to the critical threshold of 50%. The green zone indicates regions where the model’s predictions align correctly with the true classifications— predictions of LVEF less than 50% that are indeed below 50% (lower left) and predictions above 50% that are actually above 50% (upper right). Conversely, the orange zone indicates regions of misclassification— predictions above 50% for true values below 50% (lower right) and vice versa (upper left). Central to the plot is a highlighted square around the 50% line, visually representing the area of uncertainty where the model’s predictions are close to the threshold, encapsulating the challenge of near-threshold predictions. This zone of uncertainty underscores the difficulty in achieving precise classifications around the 50% cutoff point, which is critical for clinical decision-making based on LVEF values.

**Fig 5.**
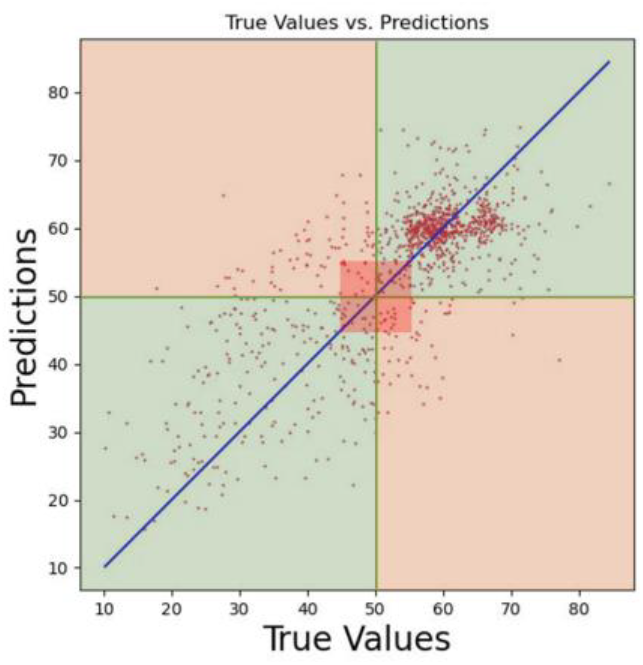
Scatter plot of true vs. predicted LVEF values by the regression model, illustrating classification accuracy relative to the 50% threshold. The green area represents correct classifications, while the orange area signifies incorrect classifications. The highlighted square around the 50% line delineates the zone of uncertainty, emphasizing the model’s challenge in making near-threshold predictions.

The study suggests that future research could focus on developing advanced classification models to identify videos with poor image quality, arrhythmias, and heart rate variations. This would improve the reliability of automated assessments by reducing the impact of the issues mentioned earlier on model predictions, thereby enhancing the accuracy of ejection fraction prediction.

## METHOD

### Data management

The study used a dataset of 10,030 apical-4-chamber echocardiography videos from patients at Stanford University Hospital between 2016 and 2018 ^38^. The data was meticulously preprocessed to ensure integrity and uniformity, including cropping and masking operations. The videos were then down-sampled to a uniform resolution of 112×112 pixels using cubic interpolation, ensuring the quality of the visual data and compatibility with the analytical framework. This dataset is crucial for understanding cardiac function representations in full resting echocardiogram studies. The dataset was divided into test, validation, and training sets, with 1,277, 1,288, and 7,462 videos in each set. The histogram in **Fig 6**a visually represents the EF values in the training set, showcasing the range from 6.90 to 96.96. The histogram shows a dataset’s imbalanced distribution of ejection fraction values, with a significant concentration in the 55% to 70% range. Consequently, the pattern and spread of EFs around the line indicate how the points in the 55% and 70% ranges are closely scattered around a diagonal line (**Fig 3**a). This imbalance can affect the performance of predictive models trained on this data, potentially leading to bias toward predicting values in the most common range. Additionally, a scatter plot was included to illustrate the spread of ejection fraction values within the training dataset (**Fig 6c**, d).

**Fig 6.**
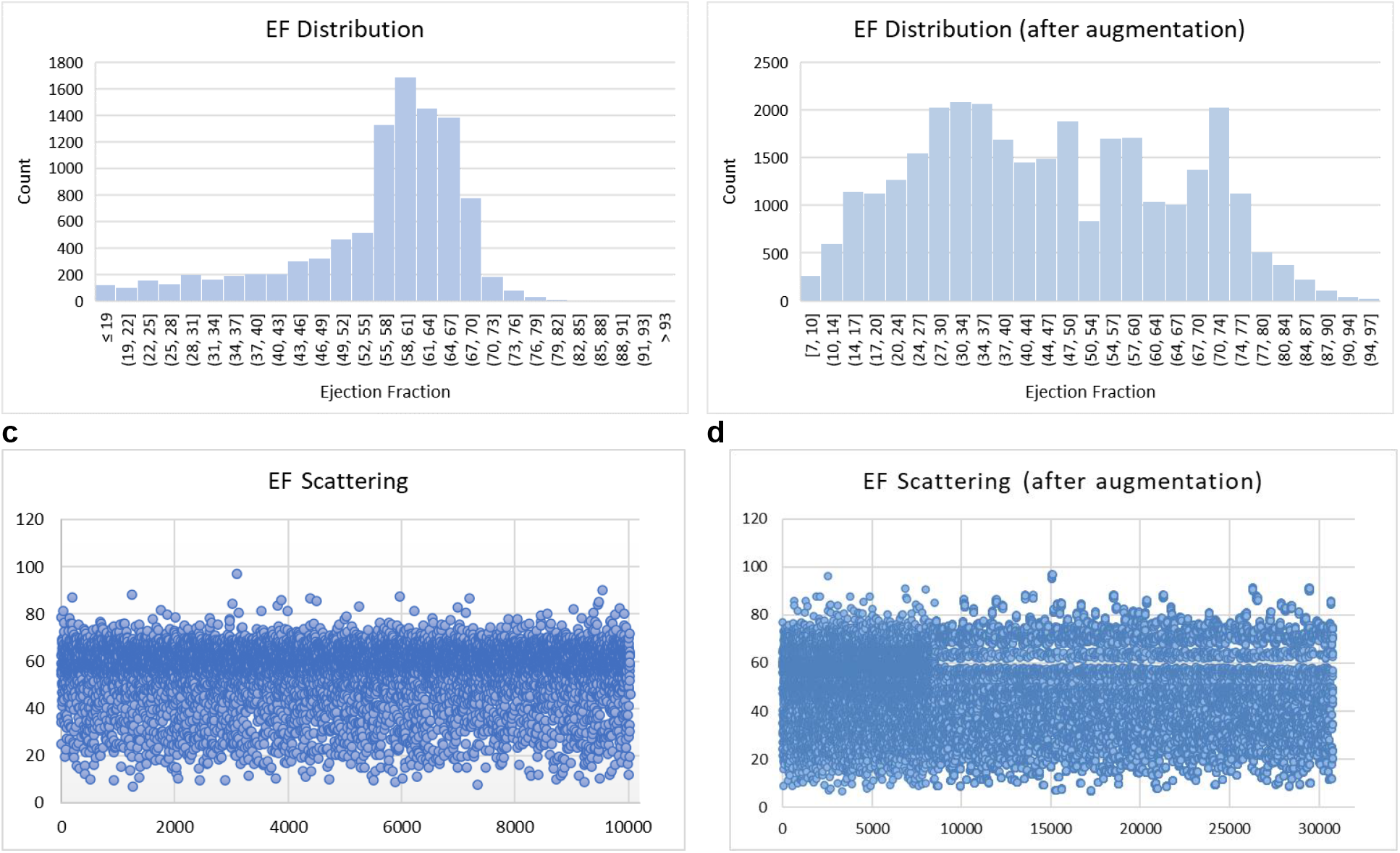
Comparative distribution of EF values in the training dataset before and after data augmentation and down-sampling techniques. The initial dataset (**a**) consisted of 7462 samples, while the augmented dataset (**b**) expanded to 30787 samples, illustrating the effect of augmentation and balancing strategies on the EF value distribution. **c** and **d** represent the spread of EF before and after augmentation in the training dataset.

In the initial examination of our training dataset, we identified a skewed distribution of EF values, which threatened to bias our predictive model towards the more common EF ranges, thereby impairing its generalizability. We first addressed the variability in frame counts to ensure uniformity in video clip length. Videos with fewer than 32 frames were lengthened by padding the last frame, whereas for videos with fewer than 64 frames, we employed 32 random samples to standardize their length. For videos containing 64 frames or more, we generated 32-frame echocardiogram clips by sampling every second frame. This preprocessing protocol was applied to all videos to create a consistent structure for subsequent steps. Following this standardization, we specifically targeted the underrepresented EF values for augmentation. For videos with an excess of 64 frames, we generated two distinct clips with variable starting points by sampling every other frame, effectively doubling the representation of these EF ranges. This augmentation, performed prior to any down-sampling, was crucial in addressing the initial data imbalance. In the subsequent phase, we down-sampled the overrepresented EF values to balance the dataset. Later, the underrepresented values are applied to each frame through a series of image transformations, including rotation, zoom, shift, and shear. A random factor between 0.99 and 1.01 also changed the EF value for each augmented video. This was done to maintain physiological plausibility and add a realistic range. The histogram in **Fig 6b** visually represents the EF values in the training set after augmentation.

### Preprocessing

Accurately assessing cardiac function using echocardiograms is crucial to minimize noise and ensure high-quality data for accurate interpretation. To address this, we developed a comprehensive preprocessing pipeline that enhances the interpretability of echocardiogram frames.

The preprocessing method starts with 32 echocardiogram frames with a 52×52 pixel resolution. The median frame is calculated by determining the median value of each pixel location across all 32 frames (temporal dimension), resulting in a singular 52×52 matrix. Then, a frame-wise multiplication operation is performed on each original video frame, resulting in a transformed video with identical dimensions but modified pixel values by multiplying the corresponding median pixel values. This meticulous operation is performed for all 32 frames in the sequence. Subsequently, histogram equalization was applied to each frame to adjust contrast and improve the visibility of cardiac structures, followed by a median blur filter with a 3×3 pixel mask to reduce noise while preserving essential anatomical details. **Fig 7** compares the first 10 frames of the original video and their preprocessed counterparts, showing the significant improvements.

**Fig 7.**
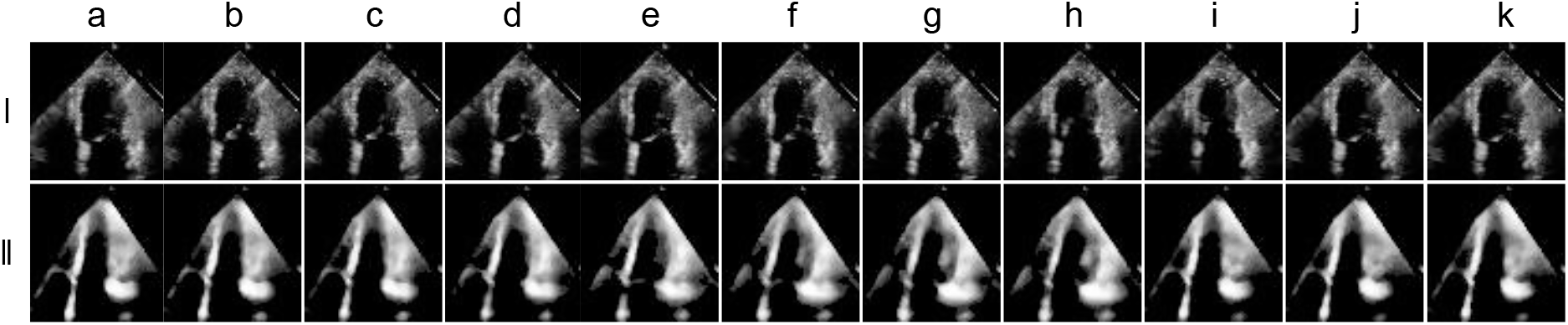
Sequential visualization of echocardiogram frame preprocessing. The top row (a-k) displays the first 11 original frames from the echocardiogram video, demonstrating the raw imaging data. After applying our preprocessing steps, the bottom row (a-k) illustrates the corresponding frames: median frame calculation, frame-wise multiplication, histogram equalization, and median blur filtering. The processed frames reveal a marked enhancement in the definition and contrast of cardiac structures, providing a clear visual distinction from the original frames and underscoring the efficacy of the preprocessing technique.

### Video-based transformer model and training

The Vision Transformer (ViT) is a pure-transformer architecture that has outperformed convolutional neural networks in image classification, offering a competitive alternative to the widely used convolutional neural networks in computer vision ^34,39^. The ViViT architecture, inspired by the ViT, provides a new approach to video classification. It uses transformer-based models, leveraging attention-based mechanisms to model long-range contextual relationships in video content. This innovative approach offers a strategic alternative to conventional 3D CNNs ^40^ and RNNs^41^, allowing for more accurate and efficient video classification.

Even though ViViT is an efficient video classification model, we trained the ViViT from scratch to directly regress the LVEF from echocardiogram videos. The model performed self-attention, computed on a sequence of spatio-temporal patches we extracted from the echocardiogram videos. We initially replaced the final layer of the classifier head, intended to output various classes, with a new layer designed to produce a single, continuous output. There is only one output unit in this new layer and no activation function. The tubelet embedding of the echocardiogram frames feeds the model with nonoverlapping spatiotemporal information.

The model is structured around a sequence of <u>ten</u> transformer layers. Each layer consists of <u>twelve</u> heads. The token size (model dimension) was set to *d* = 512. The hidden size of multi-layer perceptron (MLP) was <u>768</u>. The output of the tokens is then transformed into a regression prediction via an MLP as non-linearity in the three hidden layers of 512, 128, and 64. **Fig 8** illustrates the layered structure of our model.

**Fig 8.**
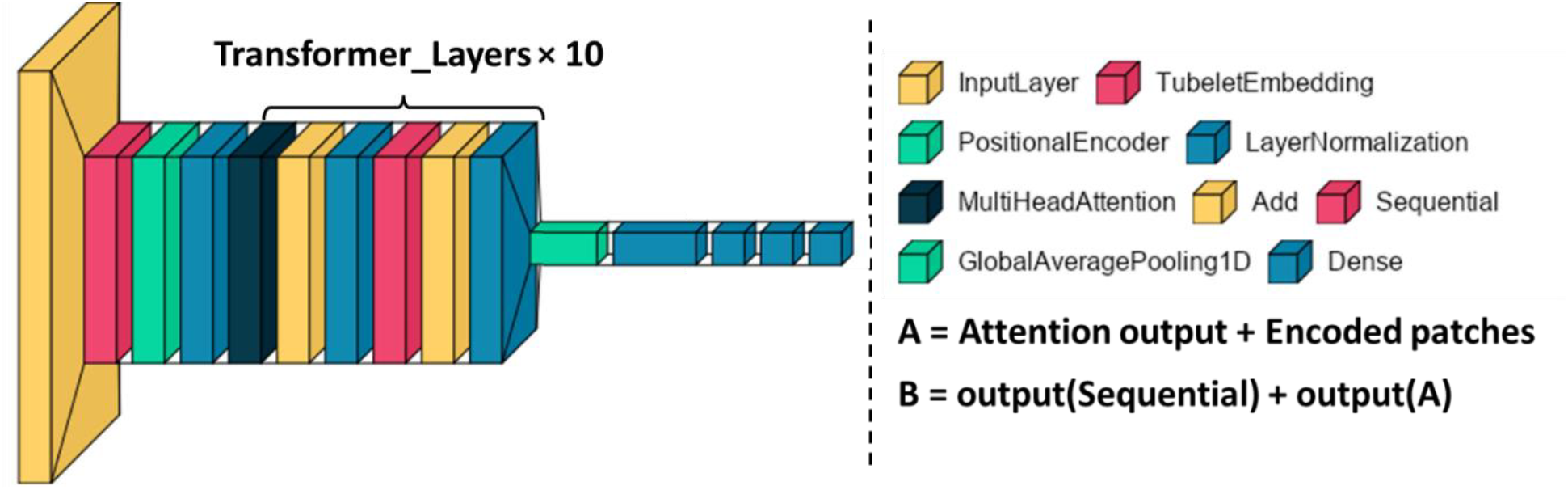
The layered structure of a neural network model. The diagram showcases the arrangement of various layers, including multi-head attention, encoder, and dense layers, illustrating the flow of information within the model.

## Data Availability

The EchoNet-Dynamic dataset was used in this project. It is a public dataset of de-identified echocardiogram videos found at https://echonet.github.io/dynamic/

## ACKNOWLEDGEMENTS

Three National Institutes of Health grants supported this work: R01HL145753, R01HL145753-01S1, and R01HL145753-03S1; in addition, the work was supported by LSUHSC-S CCDS Finish Line Award, COVID-19 Research Award, and LARC Research Award to MSB; Jane Cheever Powell Foundation for Cardiovascular Research Related to Gender and Isolation, LSUHS to SRB; and Institutional Development Award (IDeA) from the National Institutes of General Medical Sciences of the NIH under grant number P20GM121307 and R01HL149264 to CGK.

## Author Contributions Statement

Initial study concept and design: MANB, TA. Acquisition of data: TA. Model training: TA, SA. Analysis and interpretation of data: TA, SA. Drafting of the paper: TA. Critical revision of the manuscript for important intellectual content: MANB. Statistical analysis: TA, SA, MANB. Reading, editing, and approving the paper: All the authors.

## Competing interests

The authors declare no competing interests.

## Ethical Approval and Consent to participate

Not applicable.

## Replication of results

The codes and data used are available on request to enable the method proposed in the manuscript to be replicated by readers.

## Notes

### Competing Interest Statement

The authors have declared no competing interest.

### Author Declarations

We have used the public dataset, and as a result, the IRB was waived.

